# EARLY OUTPATIENT TREATMENT OF COVID-19: A RETROSPECTIVE ANALYSIS OF 392 CASES IN ITALY

**DOI:** 10.1101/2022.04.04.22273356

**Authors:** Marco Cosentino, Veronica Vernocchi, Stefano Martini, Franca Marino, Barbara Allasino, Maria Balzola, Fabio Burigana, Alberto Dallari, Carlo Servo Florio Pagano, Antonio Palma, Mauro Rango, the IppocrateOrg Association Working Group for the Early Outpatient Treatment of COVID-19

**Author notes:** **Corresponding Authors:** Marco Cosentino, MD PhD, Center for Research in Medical Pharmacology, University of Insubria, Via Monte Generoso n. 71, 21100 Varese, VA, Italy, Veronica Vernocchi, MD PhD, IppocrateOrg Association, Via Canova 15, 6900 Lugano, Switzerland. these authors provided equal contributions to the study. the list of working group members is included in Appendix 1.

## Abstract

**Introduction:** The pandemic of severe acute respiratory syndrome (SARS)-coronavirus-2 (CoV-2) disease 2019 (COVID-19) was declared in march 2020. Knowledge of COVID-19 pathophysiology soon provided a strong rationale for the early use of anti-inflammatory, antiplatelet and anticoagulant drugs, however the evidence was only slowly and partially incorporated into institutional guidelines. Unmet needs of COVID-19 outpatients were soon taken care of by networks of physicians and researchers, using pharmacotherapeutic approaches based on the best available experiences.

**Methods:** Observational retrospective study investigating characteristics, management and outcomes in COVID-19 patients taken care of in Italy by physicians volunteering within the IppocrateOrg Association, one of the main international assistance networks, between 1^st^ november 2020 and 31^st^ march 2021.

**Results:** Ten doctors took part in the study and provided data about 392 consecutive COVID-19 patients. Patients’ mean age was 48,5 years (range: 0,5-97). They were 51,3% females and were taken care of when in COVID-19 stage 0 (15,6%), 1 (50,0%), 2a (28,8%), 2b (5,6%). Many patients were overweight (26%) or obese (11,5%), with chronic comorbidities (34,9%), mainly cardiovascular (23%) and metabolic (13,3%). Drugs most frequently prescribed included: vitamins and supplements (98,7%), aspirin (66,1%), antibiotics (62%), glucocorticoids (41,8%), hydroxychloroquine (29,6%), enoxaparin (28,6%), colchicine (8,9%), oxygen therapy (6,9%), ivermectin (2,8%). Hospitalization occurred in 5,8% of total cases, mainly in patients taken care of when in stage 2b (27,3%). Altogether, 390 patients (99,6%) recovered, one patient (0,2%) was lost at follow up, and one patient (0,2%) died after hospitalization. One doctor reported one grade 1 adverse drug reaction (ADR) (transient or mild discomfort), and 3 doctors reported in total 8 grade 2 ADR (mild to moderate limitation in activity).

**Conclusions:** This is the first study describing attitudes and behaviors of physicians caring for COVID-19 outpatients, and the effectiveness and safety of COVID-19 early treatment in the real world. COVID-19 lethality in our cohort was 0,2%, while the overall COVID-19 lethality in Italy in the same period was between 3% and 3,8%. The use of individual drugs and drug combinations described in this study appears therefore effective and safe, as indicated by the few and mild ADR reported. Present evidence should be carefully considered by physicians caring for COVID-19 patients as well as by political decision makers managing the current global crisis.

## INTRODUCTION

An outbreak of pneumonia due to the novel severe acute respiratory syndrome (SARS)- coronavirus-2 (CoV-2) emerged in December 2019 in the central Chinese province of Hubei (Zhou et al., 2020), and quickly spread around the world with apparently high contagiousness and letality, and coronavirus disease 2019 (COVID-19) was eventually declared a pandemic by the World Health Organization in March 2020 (WHO, 2020). By that time, according to data from Istituto Superiore di Sanità, Italy already had 12.462 confirmed cases and 827 deaths, with a case fatality rate of 6.64%. Only China had recorded more deaths due to this COVID-19 outbreak (Remuzzi and Remuzzi, 2020).

The initial approach of the Italian Health Authorities to the management of SARS-CoV-2 spreading and COVID-19 emergence was based essentially on social distancing and isolation as well as on virological testing of patients and subsequent tracing and quarantine of asymptomatic contacts. Even autopsies of subjects died because of COVID-19 were discouraged, due to the fear for spreading of the virus (Ministero della Salute, 2020). Luckily, despite such recommendations seminal studies were performed leading to detailed description of the predominant pattern of lung inflammatory lesions in patients with COVID-19, including diffuse alveolar damage, hyaline membrane formation and pneumocyte atypical hyperplasia, and frequent extensive coagulopathy (Carsana et al., 2020), thus providing a strong rationale for the early use of anti-inflammatory, antiplatelet and anticoagulant drugs to treat COVID-19.

Unfortunately, the evidence was only partially incorporated into the various versions of the Italian Health Authorities guidelines for the early treatment of patients with SARS-CoV-2 infection, up to the latest ones (Ministero della Salute, 2021), which still recommend just symptomatic treatment of high body temperature with paracetamol and possibly with non-steroidal anti-inflammatory drugs (NSAID), warning against the extensive use of glucocorticoids and low molecular weight heparins, and advising against antibiotics, hydroxychloroquine and other antivirals. At some point, the Italian drug agency AIFA even denied authorization to the off label use of hydroxychloroquine in COVID-19 (AIFA, 2020), a decision conflicting with the right of doctors to prescribe therapeutics in the best interest of their individual patients, a doctors’ exclusive prerogative guaranteed by law.

Against this context, IppocrateOrg Association (https://ippocrateorg.org/en/), an international network joining together physicians, researchers, health and social workers, developed evidence-based guidelines for the early outpatient treatment of COVID-19, based on international experiences gained by taking care of COVID-19 from the very beginning in january 2020, on the valuable experiences of clinicians, pneumologists and infectious disease specialists who had been taking care for decades of similar infectious disease leading to interstitial lung disease, as well as by 3 taking advantage of the empirical experience of Italian doctors facing the COVID-19 epidemic in Italy. The resulting guidelines are listed in **Supplementary Tables 1-4**, and are periodically updated through regular meetings where doctors volunteering within the assistance network share and discuss their experience together with colleagues and leading experts, in the light of novel and emerging medical and scientific evidence. Such guidelines represent a non-binding reference for doctors volunteering within the assistance network. Drugs, vitamins and supplements are listed together with their suggested dosages, warnings and contraindications, and recommended according to the main COVID-19 stages (Siddiqi and Mehra, 2020; Turk et al., 2020; Cordon-Cardo et al., 2020): stage 0 (asymptomatic with positive swab), stage 1 (symptomatic without signs of lung disease), stage 2a (symptomatic with lung disease), and stage 2b (symptomatic with lung disease and desaturation). Doctors ultimately decide which drugs and drug combinations are in the best interest of the patient, which are the appropriate doses and therapeutic regimens, taking into account age, COVID-19 stage and severity, presence of relevant comorbidities and/or other risk factors. (**Supplementary Tables 1-4**).

The present study represents the first attempt to provide a comprehensive description and analysis of patients with COVID-19 taken care of by physicians volunteering within the assistance network promoted and supported by the IppocrateOrg Association. The methods section provides a detailed description of IppocrateOrg Association network history, organization and procedures. Reported results deal with the care and the outcomes of nearly 400 COVID-19 patients taken care of in Italy from 1^st^ november 2020 to 31^st^ march 2021, thus during the “second wave” of COVID-19. Outcomes are assessed in terms of recovery, without or with sequelae, hospitalization, and death.

## METHODS

### Setting

IppocrateOrg in an international association, based in Switzerland, that gathers an international network of physicians, researchers, health & social workers and ordinary people. It was founded by Mauro Rango, who – after graduating in Human Rights in Padua (Italy) – has been working for more than twenty years in the field of rights to health and access to care. The leading event to the foundation of the IppocrateOrg initiative, was a Whatsapp message sent by Mauro Rango in May 2020 to 40 Italian friends to tell them that cures for COVID-19 actually existed and that they could be accessed by everyone. The message became immediately viral and thousands of people started to contact him. IppocrateOrg was thereafter informally constituted as a Whatsapp group of hundreds of volunteers, principally doctors but also citizens, which all had the same view and mission: to guarantee the right to health and cures to every citizen. IppocrateOrg then established connections with many countries worldwide and in breaking-record time created an international network of doctors, researchers and experts in the health field, who started sharing and comparing studies, results and clinical experience on the efficacy of COVID-19 therapies and protocols. The team of medical volunteers strengthened with new members, thus making it possible to cope with the numerous requests for assistance and to establish working groups and multidisciplinary approaches. In October 2020, IppocrateOrg instituted a service of medical teleassistance, that was coordinated and managed by volunteers and doctors, in order to better manage the “second wave”. IppocrateOrg doctors developed specific approaches for the prevention and the early treatment of COVID-19. Soon it became evident that it was extremely important to administer the treatment in a timely manner, within the first few days from the beginning of symptoms. Throughout the course of the “second wave”, thousands of people were given support and were cured by doctors on call, through teleassistance. In March 2021, the book “Guarire il COVID a casa” was written by Mauro Rango and the group of volunteer doctors (Rango, 2021), and published by IppocrateOrg becoming the first manual for specific home therapies. Thousands of copies were sold and distributed by general practitioners, pharmacists and by the very same patients that were cured. In June 2021, the IppocrateOrg Association was eventually founded in Lugano, Switzerland.

### IppocrateOrg Association recommended assistance procedures

Assistance procedures consist of the following steps: “1. The patient contacts the IppocrateOrg Association and in a few hours a physician calls back the patient and, wherever necessary and possible, the physician goes to the patient’s home. 2. When the symptoms are mild, the patient is young and there are no previous pathologies, the doctor prescribes over-the-counter drugs that are readily available in any pharmacy. 3. In the event that patient’s conditions are serious, the patient is elderly or there are previous pathologies, the physician collects an accurate medical history and checks the saturation value, thereafter proposing prescription drugs at appropriate doses according to the anamnestic information. Procedures are also available on the web (https://ippocrateorg.org/assistenza-999/) and can be accessed by anyone seeking help. The service is completely free to users.” (Rango, 2021).

### Study procedures

We performed an observational retrospective study to analyze characteristics, management and outcomes of a sample of COVID-19 patients taken care of by physicians volunteering within the assistance network promoted and supported by IppocrateOrg Association. The observation period was chosen from 1^st^ november 2020, thus one month after the institution of the IppocrateOrg Association service of medical teleassistance, to allow for an initial run in period, to 31^st^ march 2021, in order to include the bulk of the second wave of COVID-19.

All the doctors volunteering during the selected period were invited to provide the following information about their COVID-19 patients: disease stage when the patient was taken care of (according to the modified staging system based on Siddiqi and Mehra (2020) and Turk et al. (2020)), demographic (age, sex) and anthropometric data (weight and height, or at least indication of under/normal/overweight or obesity), chronic comorbidities, outcome of the nasopharyngeal swab, prescribed drugs (vitamins/supplements, aspirin, antibiotics, hydroxychloroquine, ivermectin, colchicine, enoxaparin, glucocorticoids, oxygen therapy). All the information had been previously collected by physicians as routinary part of their professional activity and was provided in anonymized form. The outcomes of the study were hospitalization, recovery, without or with sequelae, or death.

### Questionnaire for participating doctors

To gain more insight into the attitudes of individual doctors concerning the choice and use of drugs included in the IppocrateOrg Association guidelines for the early outpatient treatment of COVID-19, we asked participating doctors to fill a short questionnaire investigating their propensity to use each of the drugs in the various stages of COVID-19, and the relevant factors which eventually affected their decision (**Supplementary material – Questionnaire for physicians**). A section of the questionnaire was dedicated to collect information on any suspect adverse drug reactions (ADR) observed after administration of anti-COVID-19 treatments.

### Statistical analysis

Data were collected by means of an electronic datasheet (MS Excel) and analyzed by descriptive statistics. Differences between continuous variables were assessed via the two-tailed Student’s *t* test for independent samples, while differences between categorical variables were assessed via the Fisher’s exact test. The observed significance level of the tests (P) was taken at the 5% level (0,05) or less. In any case, this value is only meant to discriminate between large correlations in the sample and is not intended to have proper inferential meaning. Answers provided by participating doctors about their attitudes towards the use of drugs in COVID-19 and their experiences with any suspect adverse drug reactions are expressed as % of responding doctors weighted for the number of COVID-19 patients included in the present study that each doctor cared for.

## RESULTS

### Characteristics of COVID-19 patients

Out of 70 doctors volunteering during the selected period, 10 (14,3%) took part in the study and provided data about a total of 392 consecutive COVID-19 patients. The number of patients taken care of was 13 (3,3% of total patients) in november 2020, 24 (6,1%) in december 2020, 51 (13%) in january 2021, 90 (23%) in February 2021, and 214 (54,6%) in march 2021. Characteristics of patients according to disease stage when patients began to be taken care of are shown in **Table 1**.

**Table 1.** Characteristics of COVID-19 patients included in the study according to disease stage when patients began to be taken care of.

|  | **COVID-19 stage** |  |  |  | **Total** |
| --- | --- | --- | --- | --- | --- |
|  | **0** | **1** | **2a** | **2b** |  |
| **n** (% of total) | 61 (15,6) | 196 (50,0) | 113 (28,8) | 22 (5,6) | 392 (100) |
| **days since symptoms began**<br>mean (min-max) | 3,3 (0-7) | 4 (0-20) | 5,5 (1-20) | 5 (1-10) | 4,4 (0-20) |
| *missing (n, % of total)* | *0 (0)* | *3 (1,5)* | *0 (0)* | *0 (0)* | *3 (0,8)* |
| **F/M**<br>(% F) | 27/34<br>(44,3) | 110/86<br>(56,1) | 58/55<br>(51,3) | 6/16<br>(27,3) | 201/191<br>(51,3) |
| **age (years)**<br>mean (min-max) | 39,2<br>(0,5-89) | 47,7<br>(1,4-97) | 51,3<br>(13-85) | 66,5<br>(44-90) | 48,5<br>(0,5-97) |
| *age (n, % of total)* |  |  |  |  |  |
| *<18* | *12 (19,7)* | *10 (5,1)* | *2 (1,8)* | *0 (0)* | *24 (6,2)* |
| *18-40* | *17 (27,9)* | *46 (23,5)* | *23 (20,4)* | *0 (0)* | *86 (22,0)* |
| *41-65* | *26 (42,6)* | *112 (57,1)* | *65 (57,5)* | *7 (31,8)* | *210 (53,7)* |
| *66-75* | *2 (3,3)* | *14 (7,1)* | *10 (8,8)* | *5 (22,7)* | *31 (7,9)* |
| *>75* | *3 (4,9)* | *8 (4,1)* | *9 (8,0)* | *10 (45,5)* | *29 (7,4)* |
| *missing* | *1 (1,6)* | *6 (3,1)* | *4 (3,5)* | *0 (0)* | *11 (2,8)* |
| **weight status**<br>n (% of total) |  |  |  |  |  |
| underweight<br>(BMI <18,5) | 3 (4,9) | 7 (3,6) | 4 (3,5) | 0 (0) | 14 (3,6) |
| normal weight<br>(BMI = 18,5-24,9) | 21 (34,4) | 77 (39,3) | 37 (32,7) | 5 (22,7) | 140 (35,7) |
| overweight<br>(BMI = 25-29,9) | 10 (16,4) | 55 (28,1) | 28 (24,8) | 9 (40,9) | 102 (26) |
| obese<br>(BMI >30) | 5 (8,2) | 17 (8,6) | 20 (17,7) | 3 (13,7) | 45 (11,5) |
| *missing* | *22 (36,1)* | *40 (20,4)* | *24 (21,3)* | *5 (22,7)* | *91 (23,2)* |
| **chronic comorbidities** | 12 (19,7) | 62 (31,6) | 47 (41,6) | 16 (72,7) | 137 (34,9) |
| n (% of total) |  |  |  |  |  |
| cardiovascular | 5 (8,2) | 38 (19,4) | 35 (31) | 12 (54,5) | 90 (23) |
| metabolic | 5 (8,2) | 25 (12,7) | 14 (12,4) | 8 (36,4) | 52 (13,3) |
| respiratory | 0 (0) | 3 (1,5) | 3 (2,6) | 3 (13,7) | 9 (2,3) |
| oncologic | 2 (3,3) | 1 (0,5) | 4 (3,5) | 3 (13,7) | 10 (2,5) |
| neurologic | 4 (6,6) | 8 (4,1) | 5 (4,4) | 1 (4,5) | 18 (4,6) |
| autoimmune | 2 (3,3) | 13 (6,6) | 7 (6,2) | 1 (4,5) | 23 (5,9) |
| **number of chronic comorbidities** |  |  |  |  |  |
| n (% of total) |  |  |  |  |  |
| 0 | 49 (80,3) | 134 (68,4) | 66 (58,4) | 6 (27,3) | 255 (66,1) |
| 1 | 7 (11,5) | 43 (21,9) | 30 (26,5) | 7 (31,8) | 87 (22,2) |
| 2 | 4 (6,6) | 13 (6,6) | 14 (12,4) | 6 (27,3) | 37 (9,4) |
| 3 | 1 (1,6) | 5 (2,5) | 2 (1,8) | 3 (13,7) | 11 (2,8) |
| 4 | 0 (0) | 1 (0,5) | 1 (0,9) | 0 (0) | 2 (0,5) |
| **nasal swab** |  |  |  |  |  |
| n (% of total) |  |  |  |  |  |
| positive | 56 (91,8) | 175 (89,2) | 110 (97,4) | 21 (95,5) | 362 (92,3) |
| negative | 0 (0) | 7 (3,6) | 0 (0) | 0 (0) | 7 (1,8) |
| not done | 1 (1,6) | 7 (3,6) | 3 (2,6) | 1 (4,5) | 12 (3,1) |
| *missing* | *4 (6,6)* | *7 (3,6)* | *0 (0)* | *0 (0)* | *11 (2,8)* |

The number of days elapsed from when symptoms started to when patients were taken care of was higher for patients taken care of in stage 2a/stage 2b (symptomatic with lung disease without/with desaturation) in comparison to those taken care of in stage 0/stage 1 (asymptomatic/symptomatic without signs of lung disease) (on average 5-5,5 days vs 3,3-4 days, P<0,01). Patients taken care of when in stage 2b were older than those in stage 2a/stage 1 (65,5 vs 51,3-47,7 years old, P<0,01), who in turn were older than those in stage 0 (39,2 years old, P<0,01). The proportion of female patients was lower in stage 2b in comparison to all the other stages, however the difference was significant only with stage 1 (27,3% vs 56,1%, P<0,05).

Patients overweight or obese increased from 24,6% in stage 0 up to 54,6% in stage 2b (P<0,05 vs stage 0). Chronic comorbidities were mainly cardiovascular (23% of total cases), metabolic (13,3%) and autoimmune (5,9%), and their frequency increased from 19,7% in stage 0, to 31,6% in stage 1, 41,6% in stage 2a (P<0,01 vs stage 0 and stage 1), up to 72,7% in stage 2b (P<0,01 vs all the other stages). The number of chronic comorbidities was not different in patients taken care of in stage 0 (mean±SD: 1,3±0,5), stage 1 (1,4±0,7), stage 2a (1,4±0,7), and stage 2b (1,8±0,8).

A positive result of the nasopharyngeal swab was available in 92,3% of the cases and in the remaining cases COVID-19 diagnosis was based on the results of later nasopharyngeal swabs and/or on the clinical signs, symptoms and disease course (including: in 3 cases anosmia, in 1 case signs of lung disease leading to hospitalization).

### Drug prescriptions

Drugs prescribed for COVID-19 are shown in **Table 2**. Vitamins and supplements, recommended by IppocrateOrg Association guidelines since stage 0, were given to nearly all the patients taken care of.

**Table 2.** Drugs prescribed throughout the cure according to the COVID-19 stage when patients began to be taken care of. Patients who were prescribed each drug or drug category are reported as absolute numbers and as percentage (in parentheses) of total patients in each COVID-19 stage.

|  | COVID-19 stage |  |  |  | Total |
| --- | --- | --- | --- | --- | --- |
|  | 0 | 1 | 2a | 2b |  |
| **n** (% of total) | 61 (15,6) | 196 (50,0) | 113 (28,8) | 22 (5,6) | 392 (100) |
| **recommended since stage 0** |  |  |  |  |  |
| vitamins and supplements | 61 (100) | 196 (100) | 108 (95,6) | 22 (100) | 387 (98,7) |
| **recommended since stage 1** |  |  |  |  |  |
| aspirin | 16 (26,2) | 139 (70,9) | 89 (78,8) | 15 (68,2) | 259 (66,1) |
| antibiotics | 17 (27,9) | 104 (53,1) | 100 (88,5) | 22 (100) | 243 (62) |
| hydroxychloroquine | 4 (6,6) | 65 (33,2) | 38 (33,6) | 9 (40,9) | 116 (29,6) |
| ivermectin | 0 (0) | 3 (1,5) | 6 (5,3) | 2 (9,1) | 11 (2,8) |
| colchicine | 1 (1,6) | 20 (10,2) | 12 (10,6) | 2 (9,1) | 35 (8,9) |
| *≥ 1 Stage 1 drug* | 26 (42,6) | 174 (88,8) | 111 (98,2) | 22 (100) | 323 (82,4) |
| **recommended since stage 2a** |  |  |  |  |  |
| enoxaparin | 3 (4,9) | 41 (20,9) | 52 (46) | 16 (72,7) | 112 (28,6) |
| **recommended since stage 2b** |  |  |  |  |  |
| glucocorticoids | 7 (11,5) | 50 (25,5) | 85 (75,2) | 22 (100) | 164 (41,8) |
| oxygen therapy | 0 (0) | 5 (2,5) | 11 (9,7) | 11 (50) | 27 (6,9) |

Among drugs recommended since stage 1, the most prescribed were aspirin (66,1% of total patients) and antibiotics (62%), followed by hydroxychloroquine (29,6%). These drugs were increasingly prescribed throughout COVID-19 stages, from 26,2%, 27,9 and 6,6% respectively, for patients taken care of in stage 0, up to 68,2%, 100% and 40,9% in patients taken care of in stage 2b. Ivermectin and colchicine, also recommended since stage 1, were given to 10% or less of patients. Overall, one or more stage 1 drugs were increasingly prescribed from 42,6% of cases in stage 0 patients, up to 88,8% in stage 1 patients, 98,2% in stage 2a patients and 100% in stage 2b patients. Prescriptions of enoxaparin, recommended since stage 2a, were only occasional in stage 0 patients (4,9%) and thereafter increased up to 72,7% of stage 2b patients. Glucocorticoids, recommended since stage 2b, followed a similar pattern, from 11,5% in stage 0 patients up to 100% of stage 2b patients. Oxygen therapy, recommended in stage 2b only when SpO_2_ <92% in ambient air, was never given in stage 0 patients, occasionally in patients taken care of in stage 1 (2,5%) and in stage 2a (9,7%), and in one out of two stage 2b patients.

### Outcomes

Outcomes of COVID-19 patients according to disease stage when patients began to be taken care of are shown in **Table 3**. Hospitalization occurred in 25 (5,8%) cases. Hospitalization rate was 1,6% among patient taken care of when in stage 0, 4,6% in stage 1, 8% in stage 2a, and 27,3% in stage 2b (P<0,05 vs stage 2a and P<0,01 vs stage 1 and stage 0). The only hospitalized stage 0 patient was a 60 year old female of normal weight and with cancer, who finally recovered with sequelae (joint pain). Stage 1 patients who underwent hospitalization, in comparison to those who were not hospitalized, did not differ for gender or weight but were older (on average, 60 year old vs 47,1 year old, P<0,05) and with more chronic comorbidities (1,3 vs 0,4, P<0,01). On the contrary, patients taken care of when in stage 2a and stage 2b who were hospitalized did not differ, in terms of either age, gender, weight or comorbidities, from patients taken care of in the same COVID-19 stage who didn’t undergo hospitalization.

**Table 3.** Outcomes of COVID-19 patients included in the study according to disease stage when patients began to be taken care of.

|  | **COVID-19 stage** |  |  |  | **Total** |
| --- | --- | --- | --- | --- | --- |
|  | **0** | **1** | **2a** | **2b** |  |
| **n** (% of total) | 61 (15,6) | 196 (50,0) | 113 (28,8) | 22 (5,6) | 392 (100) |
| **hospitalized**<br>n (% of total) | 1 (1,6) | 9 (4,6) | 9 (8) | 6 (27,3) | 25 (5,8) |
| **recovered**<br>n (% of total) | 61 (100) | 196 (100) | 111 (98,2) | 22 (100) | 390 (99,6) |
| *without sequelae* | *50 (82)* | *173 (88,3)* | *100 (90,1)* | *19 (86,4)* | *342 (88,7)* |
| *with sequelae* | *11 (18)* | *23 (11,7)* | *11 (9,9)* | *3 (13,6)* | *48 (12,3)* |
| **deceased**<br>n (% of total) | 0 (0) | 0 (0) | 1 (0,9) | 0 (0) | 1 (0,2) |
| *missing n (% of total)* | *0 (0)* | *0 (0)* | *1 (0,9)* | *0 (0)* | *1 (0,2)* |

Altogether, 390 patients out of 392 (99,6% of total patients) recovered, in 88,7% of the cases without sequelae, one patient (0,2%) was hospitalized and lost at follow up, and one patient (0,2%) died. This patient was a 72-year old overweight male with cardiovascular disease, who had been taken care on march 6^th^ 2021, when in stage 2a of the disease, and who died after hospitalization.

### Doctors’ attitudes towards the use of drugs in COVID-19

Attitudes of doctors towards the use of drugs included in the IppocrateOrg Association guidelines for the early outpatient treatment of COVID-19, according to the various disease stages, are shown in **Figure 1**. All the respondents agree about the opportunity to use vitamins and supplements in any disease stage, always in stage 0, always or often in stage 1 and stage 2a, and always, often or sometimes in stage 2b.

**Figure 1.**
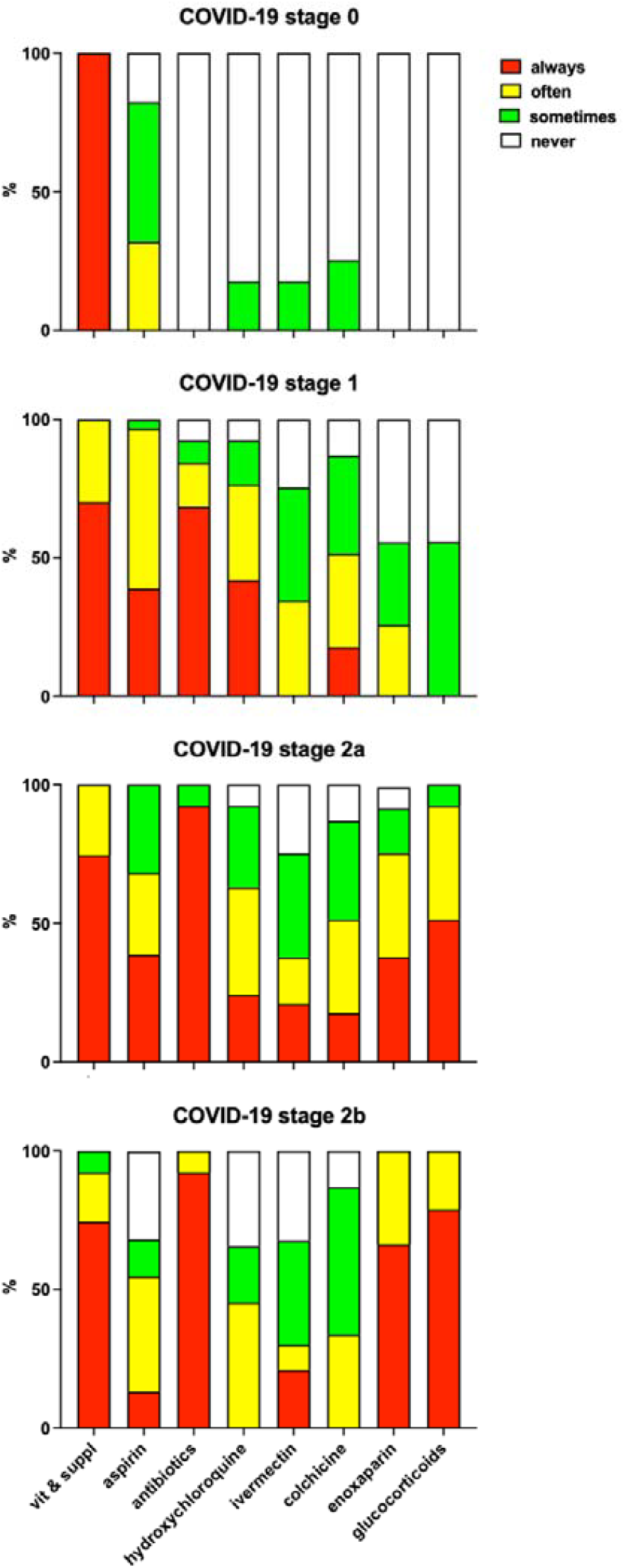
Attitudes of doctors towards the use of drugs, according to the various disease stages. Data are expressed as % of responding doctors weighted for the number of COVID-19 patients included in the present study that each doctor cared for.

In **<u>stage 0</u>**, aspirin is also considered often or sometimes in 82,3% of the cases, hydroxychloroquine, ivermectin and colchicine are considered just sometimes in 17,7-25,4%, and antibiotics, enoxaparin and glucocorticoids are never considered.

In **<u>stage 1</u>**, drugs considered always or often in more than 75% of the cases are aspirin, antibiotics and hydroxychloroquine, while colchicine is considered always or often in about 50% of the cases. Ivermectin and enoxaparin are considered often or sometimes respectively in 75,3% and 55,5% of the cases, and glucocorticoids are considered sometimes in 55,6% of the cases.

In **<u>stage 2a</u>**, antibiotics are always or often considered in 100% of the cases, and glucocorticoids, enoxaparin and aspirin are always or often considered in respectively more than 90%, more than 75% and nearly 70% of the cases. Hydroxychloroquine, colchicine and ivermectin are always or often considered in respectively in 73%, 51,4% and 37,7% of the cases.

Finally, in **<u>stage 2b</u>** antibiotics, enoxaparin and glucocorticoids are considered always or often in 100% of the cases, aspirin in 54,9%, and ivermectin in 30%. Hydroxychloroquine and colchicine are often considered in 45,3% and 33,7% of the cases.

Main reasons to consider **aspirin** in any COVID-19 stage include the occurrence of risk factors such as overweight or thrombophilia, occurrence of fever, inflammation and/or pain. Aspirin in stage 0 is considered in particular for patients who are not confident in vitamins and supplements or who cannot pay for these products.

**Antibiotics** are considered whenever fever and respiratory signs or symptoms occur, as prevention of bacterial infections. Two doctors also mention the potential antiviral activity of azithromycin. Regarding **hydroxychloroquine**, most doctors mention reasons not to prescribe this drug, including heart disease, stating that they would always consider the drug in the absence of specific contraindications. In one case, this drug is considered as an alternative whenever ivermectin is not available.

**Ivermectin** is usually perceived as highly effective in COVID-19, and preferable in comparison to hydroxychloroquine because of its safe profile. Two doctors mention that this drug is often unavailable in Italian pharmacies, and one says that during the study period (1^st^ november 2020-31^st^ march 2021) s/he didn’t know yet about the usefulness of ivermectin in COVID-19.

**Colchicine** is usually considered as an alternative to ivermectin, or in association with ivermectin and hydroxychloroquine. Remarkably, two doctors say they would choose colchicine whenever COVID-19 occurs with severe headache, and one whenever signs of pericarditis occur.

Risk of thromboembolism is the main factor leading all doctors to consider **enoxaparin** in COVID-19. Most doctors mention specific conditions such as overweight/obese or elderly subjects, hypertension, cardiac insufficiency, bedridden subjects. Three doctors also mention desaturation and respiratory symptoms/signs as reasons to consider the drug, and one says that s/he would give enoxaparin also in stage 1 whenever patients have positive anamnesis for previous thromboembolism.

Finally, **glucocorticoids** are generally considered whenever signs and symptoms, mainly fever, do not disappear after 4-5 days of treatment with other drugs and whenever SpO_2_ <95%. Careful consideration of contraindications is mentioned by most doctors.

### Perception and reporting of adverse reactions to drugs used to treat COVID-19

Perceived frequency of suspect ADR to drugs included in the IppocrateOrg Association guidelines for the early outpatient treatment of COVID-19 is shown in **Figure 2**. Drugs considered as with common ADR (frequency: 1-10%) are: antibiotics (43,2%), glucocorticoids (24,4%), hydroxychloroquine (18,7%), colchicine (17,7%), and aspirin (12,3%). Ivermectin and vitamins and supplements are associated with only uncommon ADR (frequency: <1%) and in just respectively 25,4% and 17,7% of the cases.

**Figure 2.**
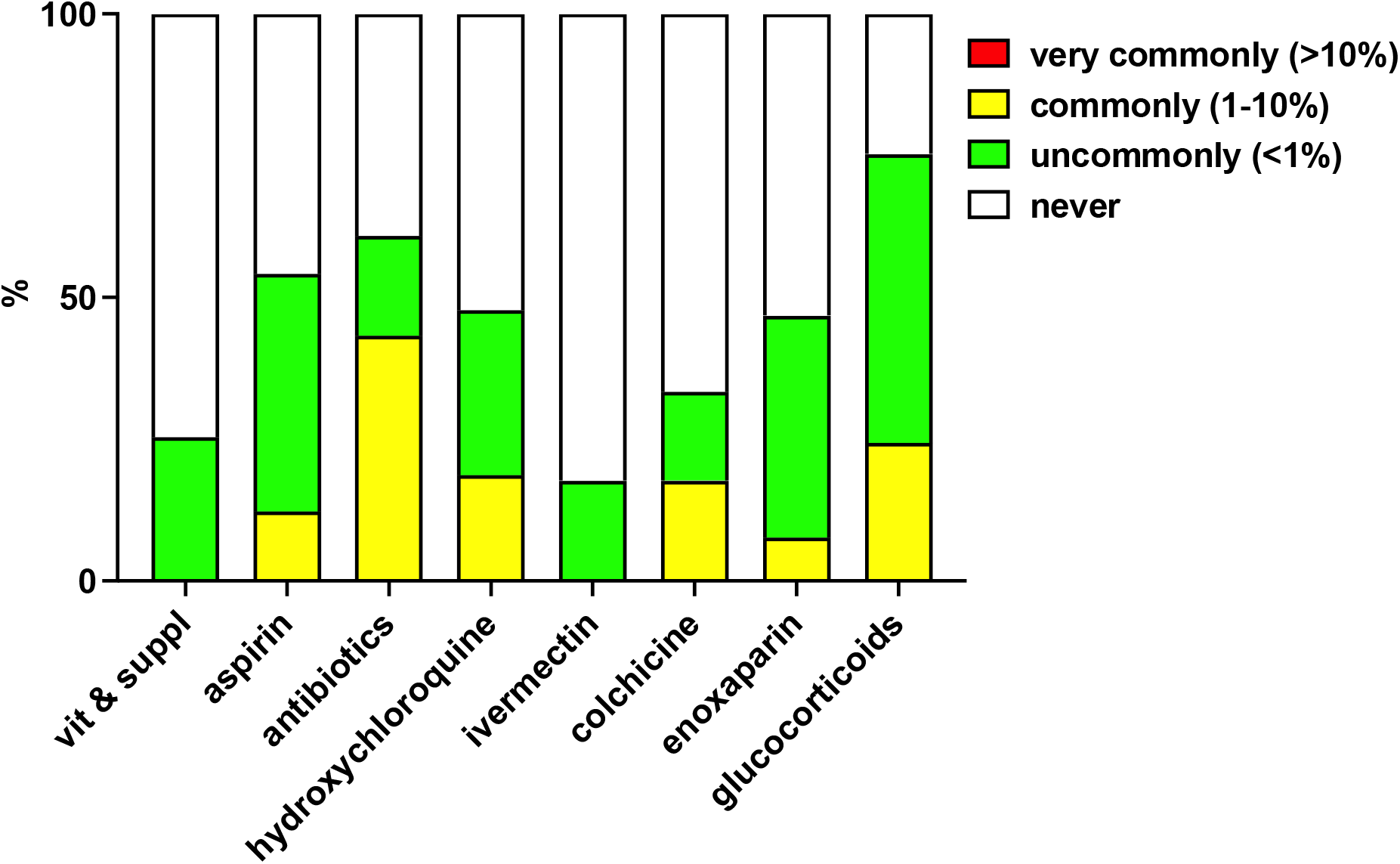
Observed frequency of suspect ADR to drugs. Data are expressed as % of responding doctors weighted for the number of COVID-19 patients included in the present study that each doctor cared for.

One doctor reported one grade 1 ADR (transient or mild discomfort (<48 hours); no medical intervention or therapy required), namely orticaria (likely due to prescribed medicines). Three doctors reported in total 8 grade 2 ADR (mild to moderate limitation in activity; some assistance may be needed; no or minimal medical intervention or therapy required), namely: gastrointestinal discomfort with aspirin, antibiotics and glucocorticoids, and hyperglycemia with glucocorticoids; atrial fibrillation and diarrhea with colchicine and hydroxychloroquine, and allergy (likely due to prescribed medicines); gastrointestinal discomfort with aspirin, diarrhea with hydroxychloroquine, persistent hiccups with glucocorticoids.

## DISCUSSION

This is the first study describing attitudes and behaviors of physicians taking care of COVID-19 outpatients, and the effectiveness and safety of early treatment of COVID-19 outpatient in the real world. Outpatient treatment of COVID-19 is still a relatively underinvestigated area and the few existing studies consist of trials of either individual therapeutics, for example hydroxychloroquine (Schwartz et al., 2021; Reis et al., 2021), colchicine (Tardif et al., 2021), azithromycin (Oldenburg et al., 2021; Hinks et al., 2021), NSAID (Guzman-Esquivel et al., 2022), corticosteroids (Clemency et al., 2022; Ezer et al., 2021; Ramakrishnan et al., 2021), antivirals (Roozbeh et al., 2021; Reis et al., 2021), antithrombotics (Connors et al., 2021), convalescent plasma (Korley et al., 2021), SARS-CoV-2 neutralizing antibodies (Gupta et al., 2021), interferons (Feld et al., 2021), sulodexide (Gonzalez-Ochoa et al., 2021), dietary supplements (Thomas et al., 2021), or fixed associations, for example hydroxychloroquine plus azithromycin (Rodrigues et al., 2021). In particular, studies so far performed in Italy include the retrospective assessment of a combination of indomethacin, low-dose aspirin, omeprazole, and a flavonoid-based food supplement, plus azithromycin, low-molecular-weight heparin, and betamethasone as needed, on the risk of hospitalization in 158 patients with early COVID-19 (Fazio et al., 2021), and a retrospective matched-cohort study comparing 90 patients with mild COVID-19 treated at home according to a recommendation algorithm (including NSAID, corticosteroids, anticoagulants, oxygen therapy and antibiotics) with 90 matched patients treated with the same drugs however without considering the algorithm (Suter et al., 2021). The pathophysiology of SARS-CoV-2 infection is however complex and requires highly individualized therapeutic approaches, taking into account individual risk factors such as age, sex and comorbidities, as well as the opportunity to combine direct antiviral therapy with anti-inflammatory and immunomodulating drugs, and antiplatelet/antithrombotic drugs (McCullough et al., 2021). For example, the I-MASK+ Early Outpatient Treatment Protocol for COVID-19, developed in 2020 by the Front Line COVID-19 Critical Care Alliance (FLCCC) and thereafter updated several times (FLCCC Alliance, 2022), includes first-line agents (antivirals, such as ivermectin and hydroxychloroquine, antiseptics for nose and mouth wash, such as chlorhexidine, povidone-iodine, and cetylpyridinium chloride, anticoagulants and immunomodulators, such as aspirin, vitamin D and melatonin, vitamins and supplements), second-line agents (such as antiandrogens, fluvoxamine and monoclonal antibodies), and third-line agents (such as corticosteroids), and nevertheless the document makes clear that treatment for an individual patient should rely on physician’s judgement, and that COVID-19 is a serious disease whose outcome depends on numerous factors including pre-existing conditions and timing of treatment. It is therefore of paramount importance to investigate how physicians manage COVID-19 patients in the real world and the effectiveness and safety of their choices. It is remarkable that physicians volunteering within the IppocrateOrg Association assistance network and participating in our survey chose drugs and drug combinations autonomously and taking into account only the best interest of the patient. Indeed, the guidelines developed and continuously updated by IppocrateOrg Association represent just a non-binding reference based on the best available scientific and clinical evidence and aimed at supporting physicians choices in individual cases (**Supplementary Tables 1-4**), and the present study is thus the first genuine attempt to provide a detailed description of early COVID-19 outpatient treatment in the real world.

The study period ranged from 1^st^ november 2020 to 31^st^ march 2021, therefore during the second wave of infections in Italy. Collected data include information on 392 COVID-19 patients, taken care of when in all the different COVID-19 stages, including 28,8% when in stage 2, thus symptomatic and with lung disease, and 5,6% when in stage 2b, thus with lung disease and desaturation. The main result of the study is about overall mortality: indeed, only one patient died of COVID-19 despite the management provided by the caring physician, therefore crude COVID-19 lethality in the present cohort of patients is 0,2%. According to official data COVID-19 lethality in Italy, standardized according to age and sex, was 6,6% during the first wave of infection from february to may 2020, 1,5% from june to september 2020, 2,4% in cases diagnosed in october 2020 (Fabiani et al., 2021), and thereafter 3% as a whole until 28 april 2021 (Task force COVID-19, 2021). According to the Wolrdometer website, overall COVID-19 lethality in Italy ranged from 11,7 to 6,9% in november 2020, from 6,6% to 4,7% in december 2020, from 4,7% to 4,2% in january 2021, from 4,1% to 3,9% in february 2021, and from 3,8% to 3,6% in march 2021 (https://www.worldometers.info/coronavirus/country/italy/). Of course, no direct comparison is possible between our cohort and whole population data, due to obvious differences in age and gender composition as well as in established COVID-19 risk factors, including overweight/obesity and chronic comorbidities. Nevertheless, the mortality rate of 0,2% (one in 392 patients) observed in our cohort is clearly much lower than anyone would expect. In particular, the patient who died was taken care of in march 2021, together with another 213 patients who eventually recovered, which leads to a crude COVID-19 lethality in patients taken care of in march 2021 of 0,5%, when the overall COVID-19 lethality in Italy was between 3% (Task force COVID-19, 2021) and 3,8% (https://www.worldometers.info/coronavirus/country/italy/).

Remarkably, the only patient deceased began to be taken care of when already in stage 2a of the disease, and his clinical profile presented several risk factors for a negative outcome, including male gender, old age (77 years), overweight (BMI: 27,7) and chronic cardiovascular disease. He was treated with vitamins and dietary supplements, aspirin, antibiotics, hydroxychloroquine and glucocorticoids, and was thereafter admitted to hospital, where he eventually died due to COVID-19. No information is available about hospital treatments. Indeed, as a whole 113 patients were taken care of when already in COVID-19 stage 2a. In this subgroup, 19 (16,8%) were more than 65 years old and 9 (8%) were more than 75, 28 (24,8%) were overweight and 20 (17,7%) were obese, 47 (41,6%) had at least one chronic comorbidity, and in particular 35 (31%) had chronic cardiovascular disease and 18 (15,1%) had two or more different chronic comorbidities.

Nevertheless, only 9 (8%) were admitted to hospital. Thus, despite the occurrence in this subgroup of several risk factors for severe COVID-19 and eventually a negative outcome, most patients recovered (111, 98,2%, since another patient was lost at follow up), suggesting the effectiveness of the pharmacotherapeutic approaches chosen by physicians. Further support to this conclusion is provided by close consideration of the subgroup or patients taken care of when already in stage 2b of the disease: most of them (15, 68,2%) were more than 65 years old and 10 (45,5%) were more than 75, 9 (40,9%) were overweight and 3 (13,7%) were obese, 16 (72,7%) had at least one chronic comorbidity, and 9 (41%) had two or more different chronic comorbidities. Nonetheless, all of them successfully recovered.

Our survey did not investigate COVID-19 course in individual patients, however some data can be taken as indirect indication of disease progression before recovery. Admission to hospital is the most obvious indicator of disease progression. In our survey, about one in 4 patients taken care of when already in stage 2b of the disease had to be admitted to hospital before recovery, however those taken care for earlier needed hospitalization only seldom (phase 2a: 8%) or very rarely (phase 1 and phase 0: 4,6% and 1,6%, respectively). Indirect indication of disease progression before recovery may also be obtained from information about drug prescriptions: Indeed, the need for oxygen therapy, which likely indicates progression up to blood desaturation (SpO_2_ <92% in ambient air) never occurred in patients taken care of when in stage 0, very rarely (2,5%) in patients taken care of in stage 1 (2,5%) and seldom when in stage 2a (9,7%), while 50% of stage 2b patients received oxygen therapy at home. Finally, according to the attitudes expressed by doctors participating in the survey towards the use of drugs in the various disease stages, it appears that antibiotics, enoxaparin and glucocorticoids would be never prescribed in stage 0 of the disease, and glucocorticoids would be never (50%) or only sometimes (50%) prescribed in stage 1. Thus the use of all these drugs in patients taken care of when in stage 0 likely indicate progression to symptomatic disease (stage 1 or beyond), and in particular the use of glucocorticoids in stage 0 or even stage 1 patients may indicate progression to stage 2. Indeed, patients taken care of when in stage 0 received antibiotics in 27,9% of the cases, enoxaparin in 4,9% of the cases and glucocorticoids in 11,5% of the cases, while patients taken care of when in stage 1 received glucocorticoids in 25,5% of the cases. As a whole, such results suggest that a minor but nevertheless not negligible fraction of COVID-19 patients progress to symptomatic and eventually severe disease before recovery. Progression may occur more frequently in the presence of specific risk factors: indeed, our results show that hospitalization rate is higher in patients taken care of when in stage 2b of the disease, and that patients taken care of when in stage 1 and eventually hospitalized were were older and with more chronic comorbidities in comparison to those who didn’t undergo hospitalization. Such observations further emphasize the need to treat COVID-19 as early as possible, and to carefully consider the presence of specific risk factors such as age and chronic comorbidities.

Doctors participating in our study were also asked to fill a short questionnaire investigating their propensity to use each specific drugs in the various stages of COVID-19, and the relevant factors which eventually affected their decision. The questionnaire also devised to collect information on any suspect ADR observed after administration of anti-COVID-19 treatments. Doctors’ answers to the questionnaire are in line with pharmacotherapeutic choices in their patients (**Table 2**). Indeed, all the respondents agree about using vitamins and supplements in any stage of the disease, to consider aspirin even in stage 0, to use in stage 1 not only stage 1 drugs but also enoxaparin and glucocorticoids (55% of respondents), and to use in stage 2 antibiotics, enoxaparin and glucocorticoids. Such attitudes fit quite well with the choices recorded in the drug utilization part of the study (Table 3): vitamins and supplements are given to all of the patients, aspirin is given to about one out of four patients taken care of when in stage 0, enoxaparin and glucocorticoids are given to most patients in stage 2, but also to one in four or five patients in stage 1. The possibility that this finding also reflects disease progression before recovery has been discussed above. Interestingly, reasons provided to choose or discard specific drugs indicated a very good knowledge of individual drug indications and possible ADR. For instance, main contraindications of hydroxychloroquine are carefully listed, conditions which recommend antibiotics or enoxaparin are well identified, as well as indications and contraindications of glucocorticoids. Doctors participating in the survey also document remarkable experience with possible ADR of drugs for COVID-19 (**Figure 2**). Observed frequency of suspect ADR is mostly rated as “never” or “uncommon” (<1%), except for aspirin, hydroxychloroquine, colchicine and enoxaparin, where ADR are rated “uncommon” or “common” (1-10%) in 15,7-41,9% of the cases and in 7,7-18,7% of the cases, respectively, and antibiotics and glucocorticoids where ADR are rated “uncommon” or “common” in 17,7-50,9% of the cases and in 23,7-43,2% of the cases. Observed ADR were however few (only 9 in the whole population) and mild (1) or mild to moderate (8). As a whole, this set of results suggest good tolerability of drugs and drug combinations used in the early COVID-19 treatment, as well as a very good knowledge of their characteristics by prescribing doctors.

Our study has many methodological limitations which we are well aware of. The main limitation is the retrospective design. Physicians were asked to provide data about patients taken care of during an emergency situation, when each of them volunteered with the overarching goal to care for patients in need and save lives. For this reason, collected data include only essential information to get an overall picture of COVID-19 patients’ clinical profile and outcomes. Much more information is needed however to describe in detail the clinical course of COVID-19 and its response to drug treatments, such as: time to recovery, drug doses and time of administration, the nature, duration and outcomes of any post-recovery sequelae, reasons for hospitalization and treatments received in hospital, etc. In addition, we lack any kind of control group, therefore results could be compared only and very cautiously with whole population official Italian data referring to about the same time periods and concerning COVID-19 lethality, but not directly compared according to age range and sex, or for different outcomes, e.g. admission to hospital. Finally, a small sample of doctors (10) took part in this first study, and nevertheless they were able to collect data about nearly 400 patients, which represents so far the largest sample of COVID-19 patients thoroughly investigated in Italy. All the participating doctors included only consecutive patients thus minimizing any possible selection bias.

In conclusion our study is the first describing attitudes and behaviors of physicians caring for COVID-19 outpatients, and the effectiveness and safety of early treatment of COVID-19 outpatient in the real world. COVID-19 lethality in our cohort of nearly 400 consecutive patients was 0,2% (only one patient died), thus the use of individual drugs and drug combinations as reported in our investigation resulted effective and safe, as also indicated by the few and mild ADR reported. The present study is anyway part of an ongoing research program with the aim to continuously collect and analyze all the information about patients taken care of. Currently, information about COVID-19 patients taken care of from april to july 2021 is being collected, while starting in august 2021 a specific procedure for the prospective collection of anonymized patients’ information has been included in the routinary activity of the IppocrateOrg Association organizational secretariat. Collected information will provide detailed information about the efficacy and safety of the personalized early outpatient treatment of COVID-19 developed and used by physicians volunteering within the network of IppocrateOrg Association. Meanwhile, we expect that the present evidence will be carefully considered by physicians taking care of their patients with COVID-19 as well as by political decision makers responsible for the management of the current global crisis.

## Data Availability

All data produced in the present study are available upon reasonable request to the authors.

## Acknowledgements

The valuable support of Dr. Rosanna Chifari Negri, Scientific Committee of IppocrateOrg Association, and of Ms. Laura Campanelli, Organizing secretariat of IppocrateOrg Association, is gratefully acknowledged. The authors would like to express their gratitude to all the physicians who volunteer within the IppocrateOrg Association collaborative network, as well as all patients who received assistance, without whom this study would not have been possible.

## Contributors

Concept and design: MC, VV, and MR. Acquisition of data: BA, MB, FB, AD, CSFP, and AP. Analysis and interpretation of data: VV, MC, SM, FM. Drafting of the manuscript: MC. Critical revision of the manuscript for important intellectual content: all authors. Statistical analysis: MC and SM. Administrative, technical or material support: VV. Data collected for the study were provided by participating physicians to IppocrateOrg Association, and are part of the information routinely collected during their professional activity. Each physician is therefore responsible for the integrity of the data s/he provided. MC, VV, SM and FM had full access to all of the anonymized data in the study and take responsibility for the integrity of the data records and the accuracy of the data analysis. All the authors accept full responsibility for the work, and controlled the decision to publish. The corresponding authors attest that all listed authors meet authorship criteria and that no others meeting the criteria have been omitted.

## Patient consent for publication

Not required.

## Ethics approval

Ethics approval has been provided by the Research Ethics Committee of the University of Insubria, Varese, Italy (project 2022_01_Cosentino).

