## Appendix 1 for "EARLY OUTPATIENT TREATMENT OF COVID-19: A RETROSPECTIVE ANALYSIS OF 392 CASES IN ITALY"

**IppocrateOrg Association Working Group for the Early Outpatient Treatment of COVID-19**

**Members:**

Barbara Allasino, Turin
Maria Balzola, Milan
Marina Basile, Catania
Rosalia Billeci, Palermo
Fabio Burigana, Trieste
Alberto Dallari, Reggio Emilia
Carlo Servo Florio Pagano, Verona
Antonio Palma, Milan
Simona Fontana, Como
Massimo Veneziano, Genova
