## Supplementary Tables 1-4 for "EARLY OUTPATIENT TREATMENT OF COVID-19: A RETROSPECTIVE ANALYSIS OF 392 CASES IN ITALY"

**Supplementary Table 1.** Vitamins and supplements recommended in COVID-19 phase 0 (asymptomatic with positive swab).

| Drug | Dosage | Contraindications/warnings | Selected references |
|---|---|---|---|
| vitamin D3 | 50,000 IU/die for 6 days (if not already taking as prophylaxis) or 10,000 IU/day for 6 days (if already taking as prophylaxis). Continue thereafter with 4,000 IU/day with a high-fat meal (i.e. at lunch). In children: 200 IU/Kg/day until a negative swab is obtained | - severe chronic renal failure<br>- hypercalcemia | Grant et al., 2020; Xu et al, 2020; Bae et al., 2020 https://c19vitamind.com/ |
| vitamin C | minimum 1 g/day, depending on formulation | - severe chronic renal failure<br>- G6PD deficiency | Bae et al., 2020; Holford et al., 2020; Colunga Biancatelli et al., 2020 https://c19vitaminc.com/ |
| zinc picolinate | 30-50 mg/day | - none | Wessels et al., 2020; Skalny et al., 2020 https://c19zinc.com/ |
| hesperidin | 100 mg/day | - none | Bellavite and Donzelli, 2020; Adhikari et al., 2021 |
| quercetin | up to 250 mg twice a day | - none | Derosa et al., 2021; Colunga Biancatelli et al., 2020 https://c19quercetin.com/ |
| lactoferrin | up to 200 mg twice a day | - none | Wang et al., 2020; Chang et al., 2020 |
| bromhexine | 8 mg three times a day. | - none | Depfenhart et al., 2020 |

| | | | |
|---|---|---|---|
| *Pelargonium sidoides* | 6 to 12 years of age: 13 mg 3 times a day for 7 days. Over 12 years of age: 20diatesi mg 3 times a day for 7 days | - haemorrhagic diathesis (for example, treatment with anticoagulants) | Brendler et al., 2021 |
| fumigations | 3 times a day | - none | |
| hydroxytyrosol and α-cyclodextrin | 2-3 oropharyngeal sprays 3-4 times a day | - none | Kiani et al., 2020; Carrouel et al., 2021 |
| vitamin A | up to 30,000 IU/day | - pregnancy | Trasino, 2020; Midha et al., 2020 |
| resveratrol | up to 1,000 mg/day | | Filardo et al., 2020 |

Important note: doctor will choose among the various options according to the profile and needs of individual patients. As a general recommendation, vitamin D3, vitamin C and zinc picolinate should be always included in any plan, also in more advanced phases of COVID-19.

**Supplementary Table 2.** Drugs recommended in COVID-19 phase 1 (symptomatic without signs of lung disease).

| Drug | Dosage | Contraindications/warnings | Selected references |
|---|---|---|---|
| acetylsalicylic acid | 100 mg/day until symptoms subside | | Bianconi et al., 2020; Cacciapuoti F, Cacciapuoti, 2021 https://c19aspirin.com/ |
| ivermectin | 0.2 mg/kg up to 20 mg total dose. If insufficient clinical response, severe disease or obesity, daily dose can be increased up to 0.4-0.6 mg/kg | | Kory et al., 2021; Marik and Kory, 2021 https://c19ivermectin.com/ |
| hydroxychloroquine | 200 mg twice a day for 7 days | - arrhythmia<br>- retinopathy<br>- G6PD deficiency | Nina and Dash, 2020; Sinha and Balayla, 2020; Million et al., 2021 https://c19hcq.com/ |
| azithromycin | 500 mg/day for 3 days, stop for 2 days, then repeat | - long Q-T Syndrome | Echeverría-Esnal et al., 2021; Million et al., 2021 |
| doxycycline (as an alternative to azithromycin) | 100 mg twice a day for 7 days | | Yates et al., 2020 |
| colchicine | 0.5 mg twice a day for 14 days | - cardiac insufficiency<br>- severe chronic renal failure | Reyes et al., 2021 https://c19colchicine.com/ |

Important note: ivermectin, hydroxychloroquine and colchicine can be prescribed only with the patient's prior written informed consent, as required by Italian law (art. 3 paragraph 2 of the Law n. 94/98, published in the Official Gazette no. 86, 14 April 1998).

**Supplementary Table 3.** Drugs recommended in COVID-19 phase 2a (symptomatic with lung disease).

| **Drug** | **Dosage** | **Contraindications/warnings** | **Selected references** |
|---|---|---|---|
| amoxicillin/clavulanic acid (consider associating azithromycin in case of bacterial superinfection) | 875 mg+125 mg 3 times a day for 8-10 days | | |
| enoxaparin | 4.000 IU/day for 10 days if <90 Kg bw<br>6.000 IU/day for 10 days if >90 Kg bw | - haemorrhagic diathesis<br>- thrombocytopaenia | Drago et al., 2020; Susen et al., 2020 |
| levodropropizine | 60 mg as needed, up to 3 times a day | | |
| acetylcysteine | 600 mg up to 3 times a day for 7 days | | Shi et al., 2020 |
| montelukast | 10 mg 2 hours after dinner for 14 days | | Aigner et al., 2020 |

Important note: doctors may consider already in this phase glucocorticoids recommended in COVID-19 phase 2b.

**Supplementary Table 4.** Drugs recommended in COVID-19 phase 2b (symptomatic with lung disease and desaturation).

| **Therapy** | **Dosage** | **Contraindications/warnings** | **Selected references** |
|---|---|---|---|
| oxygen therapy | 1-6 L/min if $SpO_2$ <92% in ambient air (*) | | |
| enoxaparin | 100 IU/Kg/12 hours | | Drago et al., 2020; Susen et al., 2020 |
| glucocorticoids | | | Alexaki and Henneicke, 2021 |
| dexamethasone | 6 mg every morning or 3 mg twice a day | - do not associate, use as alternatives<br>- use gastroprotection<br>- carefully monitor for possible hyperglycemia, elevated blood pressure, and other common glucocorticoid-associated adverse effects | |
| betamethasone | 8 mg every morning or 3 mg twice a day | | |
| methylprednisolone | 32 mg every morning or 3 mg twice a day | | |
| prednisone | 40 mg every morning or 3 mg twice a day | | |
| deflazacort | 30 mg twice a day | | |
| antibiotics | according to clinical judgement | | |

Important note: never use glucocorticoids at the beginning symptoms, wait until the end of the viremic phase. Treatment should be up to 6-7 days, thereafter taper off the drug. Prefer a single administration in the morning, consider two administrations in suffering patients. Deflazacort has a short half life and should be always given in two separate administrations.

(*) hospitalization is mandatory whenever >6 L/min needed and/or SpO2 permanently < 92%.
