## Supplemental Data 1 for "EARLY OUTPATIENT TREATMENT OF COVID-19: A RETROSPECTIVE ANALYSIS OF 392 CASES IN ITALY"

**Questionnaire for physicians**

Name ____________________________________________ date _____________________

Answer please to the following questions having in mind your attitudes and experience during the time interval included in the study (1/11/20-31/3/21).

1. How often did you prescribe **vitamins and supplements** in:

| | never | sometimes | often | always |
|---|---|---|---|---|
| **Covid-19 stage 0** | | | | |
| **Covid-19 stage 1** | | | | |
| **Covid-19 stage 2a** | | | | |
| **Covid-19 stage 2b** | | | | |

*(for each stage, only one choice allowed)*

2. How often did you prescribe **aspirin** in:

| | never | sometimes | often | always |
|---|---|---|---|---|
| **Covid-19 stage 0** | | | | |
| **Covid-19 stage 1** | | | | |
| **Covid-19 stage 2a** | | | | |
| **Covid-19 stage 2b** | | | | |

*(for each stage, only one choice allowed)*

2.1 List please criteria supporting your choice (for example presence/absence of any signs/symptoms, risk factors, indications/contraindications)

3. How often did you prescribe **antibiotics** in:

| | never | sometimes | often | always |
|---|---|---|---|---|
| **Covid-19 stage 0** | | | | |
| **Covid-19 stage 1** | | | | |
| **Covid-19 stage 2a** | | | | |
| **Covid-19 stage 2b** | | | | |

*(for each stage, only one choice allowed)*

3.1 List please criteria supporting your choice (for example presence/absence of any signs/symptoms, risk factors, indications/contraindications)

4. How often did you prescribe **hydroxychloroquine** in:

| | never | sometimes | often | always |
|---|---|---|---|---|
| **Covid-19 stage 0** | | | | |
| **Covid-19 stage 1** | | | | |
| **Covid-19 stage 2a** | | | | |
| **Covid-19 stage 2b** | | | | |

*(for each stage, only one choice allowed)*

4.1 List please criteria supporting your choice (for example presence/absence of any signs/symptoms, risk factors, indications/contraindications)

5. How often did you prescribe **ivermectin** in:

| | never | sometimes | often | always |
|---|---|---|---|---|
| **Covid-19 stage 0** | | | | |
| **Covid-19 stage 1** | | | | |
| **Covid-19 stage 2a** | | | | |
| **Covid-19 stage 2b** | | | | |

*(for each stage, only one choice allowed)*

5.1 List please criteria supporting your choice (for example presence/absence of any signs/symptoms, risk factors, indications/contraindications)

6. How often did you prescribe **colchicine** in:

| | never | sometimes | often | always |
|---|---|---|---|---|
| **Covid-19 stage 0** | | | | |
| **Covid-19 stage 1** | | | | |
| **Covid-19 stage 2a** | | | | |
| **Covid-19 stage 2b** | | | | |

*(for each stage, only one choice allowed)*

6.1 List please criteria supporting your choice (for example presence/absence of any signs/symptoms, risk factors, indications/contraindications)

7. How often did you prescribe **enoxaparin** in:

| | never | sometimes | often | always |
|---|---|---|---|---|
| **Covid-19 stage 0** | | | | |
| **Covid-19 stage 1** | | | | |
| **Covid-19 stage 2a** | | | | |
| **Covid-19 stage 2b** | | | | |

*(for each stage, only one choice allowed)*

7.1 List please criteria supporting your choice (for example presence/absence of any signs/symptoms, risk factors, indications/contraindications)

8. How often did you prescribe **glucocorticoids** in:

| | never | sometimes | often | always |
|---|---|---|---|---|
| **Covid-19 stage 0** | | | | |
| **Covid-19 stage 1** | | | | |
| **Covid-19 stage 2a** | | | | |
| **Covid-19 stage 2b** | | | | |

*(for each stage, only one choice allowed)*

8.1 List please criteria supporting your choice (for example presence/absence of any signs/symptoms, risk factors, indications/contraindications)

9. How often did you observe any suspect adverse reactions with each of the following drugs?

| | **never** | **uncommonly** | **commonly** | **very commonly** |
|---|---|---|---|---|
| | | <1% | 1-10% | >10% |
| **vitamins and supplements** | | | | |
| **aspirin** | | | | |
| **antibiotics** | | | | |
| **hydroxychloroquine** | | | | |
| **ivermectin** | | | | |
| **colchicine** | | | | |
| **enoxaparin** | | | | |
| **glucocorticoids** | | | | |

10. In case you observed any suspect adverse reactions, please indicate whether they were:

| |
|---|
| **Grade 1: Mild**<br>*transient or mild discomfort (< 48 hours); no medical intervention or therapy required* |
| **Grade 2: Moderate**<br>*mild to moderate limitation in activity; some assistance may be needed; no or minimal medical intervention or therapy required* |
| **Grade 3: Severe**<br>*marked limitation in activity, some assistance usually required; medical intervention or therapy required, hospitalizations possible* |
| **Grade 4: Life threatening**<br>*extreme limitation in activity, significant assistance required; significant medical intervention or therapy required, hospitalization or hospice care probable* |

11. In case you observed grade 2 or higher suspect adverse reactions, please briefly describe the case(s):
